# Feasibility and Preliminary Effects of a Culturally Adapted Internet-Based Mental Health Promotion Intervention for Indonesian Undergraduate Students (LifeHack: Menjadi Mahasiswa Tangguh): A Protocol for a Single-Arm, Pre–Post Feasibility Study

**DOI:** 10.64898/2026.10.27.26364572

**Authors:** Siti Raudhoh, Hari Setyowibowo, Sascha Struijs, Aulia Iskandarsyah

## Abstract

**Background:** Mental health problems are highly prevalent among university students globally and in Indonesia, yet the vast majority do not receive adequate care. Whilst existing digital mental health interventions in Indonesia have primarily targeted students with clinically elevated symptoms, no prior study has evaluated an internet-based intervention designed to promote positive mental health among Indonesian university students. LifeHack is an internet-based guided self-help intervention developed to promote positive mental health as part of the Caring Universities initiative within the WHO World Mental Health–International College Student (WMH-ICS) framework. LifeHack: Menjadi Mahasiswa Tangguh is a culturally adapted Indonesian version, developed through a systematic adaptation process guided by Bernal’s Ecological Validity Model and the Heuristic Framework for Cultural Adaptation. Objectives: This study aims to evaluate the feasibility and acceptability of the adapted intervention, including its usability, participant satisfaction, adherence, and retention. Secondarily, it will examine preliminary signals of change in resilience, positive mental health, depression and anxiety symptoms, and coping strategies.

**Methods:** This protocol describes a single-arm pre–post feasibility study involving 50 undergraduate students aged 18 years or older from Universitas Padjadjaran and Universitas Jambi, Indonesia. Participants will receive the culturally adapted LifeHack: Menjadi Mahasiswa Tangguh, a self-paced web-based guided self-help intervention supported by asynchronous personalised feedback from trained e-coaches. Participants may select modules according to their individual needs and complete the intervention over a flexible period of 1–6 weeks. Assessments will be conducted at baseline (T0), immediately following intervention completion or at the end of the six-week intervention period if participants have not previously indicated completion (T1), and three months after T1 (T2). Primary feasibility outcomes include usability, satisfaction, intervention adherence, retention, and intervention completion. Secondary outcomes include resilience, positive mental health, psychological distress, coping, and safety. Safety will be assessed through participant-reported negative effects and recorded adverse events identified through routine e-coach monitoring. Progression to a future randomised controlled trial will be evaluated using pre-specified traffic light criteria.

**Expected Outcomes:** Findings will inform the feasibility, cultural appropriateness, and preliminary effects of the adapted LifeHack intervention for Indonesian students. Results will guide potential progression to a fully powered randomised controlled trial.

**Trial Registration:** Thai Clinical Trials Registry (TCTR), TCTR20260530005—, registered on 9^th^ May 2026 and updated on 17^th^ August 2026

## Introduction

Mental health problems are common among young people worldwide. Global Burden of Disease estimates indicate that a substantial proportion of individuals aged 20–24 live with a mental disorder, approximately 16% (95% UI 13.9–19.4) (1). This pattern suggests that young adults—particularly university-aged populations—experience a higher burden of mental health problems compared to the general population, reinforcing their heightened vulnerability to conditions such as depression and anxiety (1). Consistent with this, around a third of students arrive at university with symptoms of a common mental disorder (2), and roughly 38% screen positive for at least one 12-month disorder (3). Another systematic review also reports prevalence rates of 25% for depression and 14% for suicidal ideation among university students (4). Similar patterns are observed in Indonesia, where studies during the COVID-19 pandemic reported high levels of stress (44.6%), anxiety (47.8%), and depression (18.6%) among undergraduate students (5), alongside substantial levels of psychological distress in other student populations (6).

Despite this high burden, access to adequate mental health care among university students remains limited. Although effective treatments exist, only one in five students with mental disorders receive minimally adequate treatment (7). This gap is also evident in Indonesia, where most students with mental health problems do not receive adequate care (8). In low- and middle-income countries such as Indonesia, structural constraints — including an acute shortage of mental health professionals relative to the student population — further exacerbate this gap (9). Critically, treatment-oriented approaches alone are insufficient to address population-level mental health burden: they reach only those who already present with elevated symptoms, often after significant distress has accumulated (10). What is needed alongside treatment capacity is a shift toward promotive approaches— interventions that target the general student population not merely to prevent or reduce symptoms but to actively build students’ psychological strengths and enhance positive functioning and well-being (11,12). A recent mega-analysis of 198 meta-analyses found small-to-medium effects of positive psychology interventions on well-being and psychopathology (13).

Digital mental health interventions offer a scalable and cost-effective pathway for delivering both treatment and promotive intervention at scale (14,15), with meta-analytic evidence supporting their effectiveness in enhancing well-being (16) and reducing symptoms of anxiety, depression, and stress in university populations (17). In Indonesia, a growing body of research has begun to evaluate culturally adapted digital interventions for students, including programs targeting depression (18), transdiagnostic symptoms of anxiety and depression (19), stress (20), emotional distress (21), and psychological distress (22). Across these studies, eligibility was determined by elevated symptom or distress scores, and outcomes centered on reducing depression, anxiety, stress, or distress—even where well-being was assessed (22), it remained a secondary measure within an illness-reduction framework. This literature has thus been oriented predominantly toward treatment and indicated prevention (23), designed to alleviate symptoms once distress has already emerged. This study extends this literature toward a promotive approach, one designed to build resilience and positive mental health as its primary aim, accessible to students regardless of their current symptom status. Such an approach targets a distinct outcome from the illness-reduction paradigm that has so far dominated digital mental health research in Indonesia. It complements existing treatment and indicated-prevention services by reaching students regardless of symptom level — including those without elevated symptoms — who can benefit from strengthening their psychological resources and positive mental health (12,24).

LifeHack is an internet-based intervention developed within the Caring Universities initiative — a collaborative project embedded within the World Health Organization World Mental Health– International College Student (WMH-ICS) framework in the Netherlands. The intervention was developed using a co-creation approach involving university students and mental health professionals; co-creation as a participatory design strategy leverages the experiential knowledge of end-users to enhance relevance and acceptability (25). LifeHack is a guided internet-based self-help programme grounded in cognitive behavioural techniques with an evidence base spanning multiple meta-analyses demonstrating effectiveness in improving mood and reducing stress in the general and student population (7,26–28). Its primary goal is to improve mental well-being by increasing resilience, building life skills, and strengthening positive mental health. The program was developed based on the existing empirical literature and further refined through direct collaboration with university students via interviews and co-creation, ensuring that the content and format reflect the specific needs and preferences of the student population (25). LifeHack comprises 12 modules across four thematic domains—emotional well-being, study skills, social life, and self-esteem—each taking approximately 30 minutes. Content is delivered via text, images, and infographics, with some modules including video and audio elements; each module includes evidence-based information, exercises, and homework assignments. The program is delivered via guided internet-based self-help, with asynchronous written personalized feedback provided by trained e-coaches within 48 working hours of module completion.

To extend this evidence to a new sociocultural context, the present study evaluates the feasibility and preliminary effects of *LifeHack: Menjadi Mahasiswa Tangguh—a* culturally adapted Indonesian version of the intervention. The systematic process of this cultural adaptation was guided by two complementary frameworks: Bernal’s Ecological Validity Model (29), specifying eight dimensions along which interventions may be culturally modified (language, persons, metaphors, content, concepts, goals, methods, and context); and the Heuristic Framework for Cultural Adaptation (30), which structured the adaptation across four sequential stages. Together, these frameworks ensured that surface and deep structural modifications preserved the evidence-based active ingredients of the original program. The adaptation process is detailed in the Methods section.

The present study constitutes Stage 3 and 4 (adaptation testing and refinement) of the Heuristic Framework (31), following completion of Stages 1 and 2 (information gathering and preliminary adaptation) prior to this study. Its findings will determine whether further modifications are warranted and will provide the design parameters needed for a fully powered randomised controlled trial (RCT). The study addresses the following hypotheses:

1. **H1:** The recruitment process will be considered feasible if at least 70% of eligible students who submit an Expression of Interest (EOI) enrol within the planned recruitment period.
2. **H2:** The LifeHack: Menjadi Mahasiswa Tangguh platform will demonstrate acceptable usability, with at least 70% of participants achieving a System Usability Scale (SUS) score of 68 or above.
3. **H3:** Participant adherence will be considered acceptable if at least 70% of active participants complete at least one module, click “finish”.
4. **H4:** Preliminary signals of improvement will be observed in resilience (RES) and positive mental health (MHC-SF) scores from baseline (T0) to post-intervention (T1), defined as a within-group effect size of at least Cohen’s d = 0.2 (small effect) (32).

## Methods

### Study Design

This study is an open-label, single-arm pre–post feasibility study. This non-randomised pre–post design with the primary aim of evaluating feasibility will generate evidence to inform the design and procedures of a future fully powered RCT (33). The study is best characterised as a quantitative feasibility study with embedded qualitative feedback. Its primary aim is to generate quantitative feasibility metrics—recruitment, retention, adherence, module completion, and the variability of preliminary outcome measures—while qualitative data serve a secondary, supportive role. Qualitative feedback will be collected through open-ended questions administered at the end of each module and, where participants are contacted by the researcher or an e-coach following disengagement, from the reasons given for discontinuing; these data will be analysed using qualitative content analysis to help interpret the quantitative findings and to refine the intervention content and study procedures prior to a definitive trial. In addition, preliminary effect estimates on secondary psychological outcomes will be reported to guide sample size calculations for future trials.

The reporting of this protocol follows the Standard Protocol Items: Recommendations for Interventional Trials (SPIRIT) expanded checklist 2025 (34), supplemented by relevant items from the CONSORT 2025 statement (35). The SPIRIT and CONSORT expanded checklist is provided as a supplementary file.

Three assessment time points are employed: baseline (T0), post-intervention (T1), and follow-up (T2). T1 will be administered immediately after participants indicate completion of the intervention by clicking the “Finish” button, or at the end of the six-week intervention period if they have not previously indicated completion. T2 will be administered three months after T1. The assessment schedule, following the recommended SPIRIT format, is presented in Fig 1.

### Participants and eligibility criteria

Participants will be 50 undergraduate students recruited from two public universities in Indonesia – Universitas Padjadjaran (Bandung, West Java) and Universitas Jambi (Jambi) – with a target of 25 participants from each university. Eligible participants must meet all of the following inclusion criteria:

a. Aged 18 years or older;
b. Currently enrolled as an active undergraduate student at Universitas Padjadjaran or Universitas Jambi;
c. Have access to a digital device (smartphone or computer) with reliable internet connectivity;
d. Score at or below one standard deviation above the mean (Mean + 1 SD) of the MHC-SF total score in the screening sample, calculated after the screening period closes by pooling all students screened at both universities. This criterion ensures that students with already optimal mental well-being, who would have little room for improvement (ceiling effect), are not included
e. Provide informed consent to participate.

Individuals will be excluded if they meet any of the following exclusion criteria:

a. Report a current risk of suicidal behaviour, operationally defined as: PHQ-9 item 9 score ≥ 1, AND positive endorsement of having thought about or worked out a plan to kill themselves in the past 12 months, AND reporting that this is at least somewhat likely they will act on this plan in the next 12 months;
b. Exhibit severe depressive symptoms, defined as a total PHQ-9 score of 20 or above;
c. Self-report a current diagnosis of a severe psychiatric disorder (e.g., psychotic disorder, bipolar disorder with active manic episode, or severe personality disorder requiring intensive clinical management);
d. Self-report a current diagnosis of a significant neurocognitive disorder that would impair their ability to engage with written intervention content;
e. Are anticipated to be unavailable for the full study period, including the follow-up assessment three months after T1—e.g., due to planned academic leave or extended travel.

### Recruitment

Participants will be recruited from 1-24 October 2026 through multiple strategies targeting undergraduate students at Universitas Padjadjaran and Universitas Jambi. Recruitment channels will include: (1) announcements disseminated via university lecturers and student coordinators; (2) digital posters distributed through official university social media accounts and student WhatsApp groups; and (3) recruitment invitations sent by the universities directly to students’ institutional email addresses. Recruitment materials will include a brief description of the study, its purpose, eligibility criteria, and a link to the online screening form. Recruitment and screening will run concurrently at both universities throughout this period.

The selection and modification of recruitment strategies will be guided by available resources, time constraints, and the speed of recruitment. The time taken, source of recruitment, and number of recruitment channels utilised to achieve the target sample size will be recorded as part of the feasibility evaluation.

The online screening questionnaire will include the PHQ-9 to determine eligibility, together with general demographic questions and brief items regarding current or previous mental health diagnoses, current or previous use of mental health services, including psychotherapy and psychotropic medication, and the reason for interest in the intervention. Current use of psychotherapy, psychotropic medication, or other mental health services will not constitute an exclusion criterion but will be recorded descriptively to characterize the sample and inform the interpretation of changes in psychological outcomes.

In this study the MHC-SF and PHQ-9 serve both as a screening instrument (see Recruitment) and as outcome measures administered at T0, T1, and T2. Participants who endorse PHQ-9 item 9 (score ≥ 1) will be asked two additional questions assessing whether they have thought about or worked out a plan to kill themselves in the past 12 months and how likely they are to act on this plan in the next 12 months. These additional questions reflect established distinctions between suicidal ideation, suicide plans, and suicidal behaviour used in suicide-risk assessment research (36,37). Participants will meet the predefined suicide-risk exclusion criterion if they endorse PHQ-9 item 9 (score ≥ 1), report having thought about or worked out a suicide plan in the past 12 months, and report being at least somewhat likely to act on this plan in the next 12 months. Participants can withdraw from the study at any time without penalty.

Individuals who are found ineligible due to meeting the predefined suicide-risk exclusion criterion or severe depressive symptoms will be notified via email within 48 hours of completing the screening questionnaire. They will be informed that they do not meet the current eligibility criteria and provided with written information about accessible professional mental health resources, including university counseling services and national crisis helplines (e.g., www.healing119.id or by calling 119 ext. 8).

Once the screening period closes, the MHC-SF cut-off (Mean + 1 SD) will be calculated from the full screening sample, and the resulting value will be reported. All remaining individuals will then be notified of their eligibility by email. Those whose MHC-SF score exceeds the cut-off will be thanked for their interest and informed that the study is intended for students currently experiencing less than optimal mental well-being. The number of individuals excluded for each reason will be recorded as part of the recruitment feasibility evaluation.

### Procedures

Eligible participants—those who pass the online screening—will be presented with detailed study information and an electronic informed consent form. Those who provide informed consent will immediately proceed to the baseline assessment (T0) within the same form, administered via Google Forms. Participants will be given one week to complete the consent and baseline form, with a reminder email sent to those who have not yet completed it. At the end of this period, on 1 November 2026, participants who have completed T0 will receive access credentials to the LifeHack: Menjadi Mahasiswa Tangguh platform by email, so that all participants begin the intervention on the same date. Data collected at different stages will be linked using a unique participant identification code and stored without direct personal identifiers, with access restricted to named members of the research team.

Prior to commencing the intervention modules, all participants will be invited to view a brief onboarding video. The video will cover (1) a concise overview of the study; (2) how to navigate the intervention platform; (3) the module structure and completion requirements; (4) an introduction to the e-coaches, and (5) how to contact the e-coaches and access support if needed. Viewing the onboarding video is encouraged to support engagement but is not a condition of eligibility or continued participation.

Participants will be given six weeks to engage with the intervention. Participants are required to complete at least one intervention module. Once they consider their use of the intervention complete, they will click the “finish” button to indicate completion and proceed directly to the post-intervention assessment. Ongoing monitoring of engagement will be conducted: participants who have not logged into the platform for two consecutive weeks will be contacted via email for a welfare check-in and to encourage continued engagement. Reasons for non-adherence will be recorded where provided. Any reasons given for reduced engagement or discontinuation will be recorded and, where provided, used as qualitative feedback to help interpret the feasibility findings.

Participants who have completed the minimum module requirements and indicate that they have finished the intervention will proceed to the post-intervention assessment (T1). Participants who have not indicated completion by the end of Week 6 will be invited to complete T1 at that point regardless. A follow-up assessment (T2) will be administered three months after completion of the post-intervention assessment (T1) to examine short-term maintenance of outcomes.

Participants who complete both T1 and T2 will receive compensation in the form of mobile data credit totaling IDR 150,000, distributed in two installments: IDR 100,000 upon completion of T1, and IDR 50,000 upon completion of T2. This compensation is framed as reimbursement for data (quota) costs rather than as a performance incentive. To reduce the likelihood that participation is motivated primarily by financial gain, the compensation will be disclosed only during the informed consent process and not in the recruitment materials or the study overview presented before screening.

### Intervention

#### Cultural adaptation process

LifeHack: Menjadi Mahasiswa Tangguh is a cultural adaptation—not a newly developed intervention -- of the evidence-based Caring Universities programs. The adaptation process was guided by the Heuristic Framework for Cultural Adaptation (30), operationalised through the eight dimensions of Bernal’s Ecological Validity Model (29), across four sequential stages.

Stage 1—Information Gathering: A review of the evidence base for digital mental health interventions for Indonesian university students was conducted, alongside a needs assessment of the target population. This stage established the rationale, priorities, and scope of the adaptation.

Stage 2—Preliminary Adaptation Design: The original program content was translated using a dual forward-translation and dual backward-translation procedure conducted by independent translators. Discrepancies were reconciled to produce a consolidated Indonesian version. Contextual modifications were then applied across the eight dimensions of Bernal’s Ecological Validity Model— including language, cultural metaphors, content, visual materials, and contextual references—whilst retaining the CBT-based theoretical foundation, module structure, and active ingredients of the original programs.

Stage 3—Preliminary Adaptation Testing: The adapted draft was reviewed by a supervisory panel of three licensed psychologists to evaluate conceptual consistency and cultural appropriateness. Visual materials (e.g., images and memes) were evaluated through a student survey assessing comprehensibility, relatability, and engagement value, followed by a focus group discussion to refine culturally appropriate elements. Subsequently, an expanded expert and user review involving five clinical psychologists and 25 undergraduate students evaluated functional, conceptual, and linguistic equivalence relative to the original programs. Feedback from this review informed final refinements.

Stage 4 -- Adaptation Refinement: The present feasibility study constitutes this stage. Findings will determine whether further modifications to the intervention or study procedures are warranted prior to progression to a fully powered RCT.

#### Structure and content of the adapted intervention

The adapted intervention, *LifeHack: Menjadi Mahasiswa Tangguh*, consists of 12 modules organised into four thematic domains: (1) emotional well-being, (2) study skills, (3) social life, and (4) self-esteem. The overall theoretical foundation and promotive objectives of the original LifeHack programme were retained to ensure conceptual fidelity. Module titles and theme names have been translated and contextually adapted into Indonesian, whilst the article uses English terminology throughout for consistency.

Each module is designed to take approximately 30 minutes to complete. Content is delivered via text, images, and infographics, with some modules including video and audio elements. Each module includes evidence-based psychoeducational information, structured exercises, and homework assignments. Participants engage in activities aimed at strengthening resilience-related capacities, adaptive coping strategies, and positive mental health processes. Several modules incorporate structured goal-setting exercises in which participants develop actionable plans. At the end of each module, participants complete a brief evaluation. All module content in the Indonesian version has been adapted into Indonesian, with contextual modifications applied to language, examples, and visual materials to reflect the lived experiences of Indonesian university students.

In the Indonesian adaptation, participants are required to complete a minimum of one module and indicate completion by clicking the “Finish” button at the end of the module. This minimum completion requirement is a contextual adaptation to support the feasibility evaluation, whereas the original LifeHack programme allows students to engage with the modules as standalone units based on their individual needs and preferences. Participants will retain access to the intervention during the waiting period before the T2 assessment. They may revisit previously completed modules or access any module at any time, consistent with the intervention’s intended real-world implementation as an on-demand resource that students can use whenever support for a specific topic is needed.

The intervention is delivered entirely online via a dedicated web-based platform and is supported by asynchronous written e-coaching. Participants submit responses to module exercises and receive structured written feedback from trained e-coaches within 48 working hours of submission. Coaching is guided by standardised templates and supervised by a licensed psychologist to maintain intervention fidelity. All coaching interactions are documented within the platform. A brief description and list of topics covered in each of the 12 modules is provided in Supplementary Materials 1.

In the Indonesian adaptation, e-coaching will be delivered by psychologist interns or master’s students in psychology, all of whom hold a bachelor’s degree in psychology. Psychologist interns are students enrolled in Indonesia’s Professional Psychology Program who are completing their supervised professional internship as part of their pre-licensure training to become licensed psychologists. In the Indonesian context, both psychologist interns and psychology master’s students have completed foundational training in psychology and possess competencies in applied psychological knowledge and skills. However, their role in the intervention will remain limited to e-coaching rather than psychological assessment, counseling, or treatment, to preserve the self-guided nature of the digital intervention and prevent excessive therapist-like input. E-coaches will provide asynchronous, written, personalized feedback within 48 working hours of module completion to support motivation, adherence, and understanding of module content. They will receive standardized training and ongoing supervision, and participants will receive email reminders in case of non-engagement. E-coaching fidelity will be assessed through regular review of a random sample of coaching exchanges by the supervising psychologist using a standardized fidelity checklist.

The e-coach training materials and manual will also be culturally adapted to the Indonesian context, including guidance on culturally appropriate communication, safety procedures, and responses to potential adverse or negative reactions to the intervention. Clear escalation procedures will be provided for concerns beyond the e-coach’s role, with referral to the supervising licensed psychologist when needed. E-coaches will also receive training in psychological first aid to support appropriate initial responses in situations requiring urgent assistance. These adaptations aim to strengthen participant support while maintaining the intended intensity and boundaries of the digital intervention.

### Outcomes

#### Primary Feasibility Outcomes

The primary outcomes of this feasibility study are indicators of usability, satisfaction, adherence, and retention. A predetermined traffic light system will be used to inform whether proceeding to a fully powered randomized controlled trial (RCT) is warranted. The three-tiered progression criteria require pre-specified cutoffs for a green zone (proceed without modification), amber zone (proceed with amendments), and red zone (do not proceed) (38). Consistent with current guidance for pilot and feasibility studies, these criteria will be treated as guidelines rather than mechanical rules (39,40). Accordingly, rather than basing the decision on the single worst-performing criterion, the criteria will be interpreted together and mapped to their corresponding feasibility objectives, taking into account contextual factors and the embedded qualitative feedback. On this basis, the research team— comprising a clinical psychologist, a digital health researcher, and a methodologist—will make a holistic progression decision, distinguishing whether the trial procedures are feasible (“can we?”) from whether progression is warranted (“will we?”), and the rationale for the final decision will be documented and reported transparently. As this guidance was developed primarily for randomized pilot trials, it is applied here with adaptation to the present single-arm feasibility design.

The pre-determined thresholds are presented in Table 1. Each threshold was derived from domain-specific evidence rather than applied uniformly across all criteria. Recruitment and retention thresholds were informed by published feasibility benchmarks from randomized pilot trials in psychological and digital health interventions (41–43), with additional grounding in pilot evidence from digital mental health interventions specifically targeting university student populations (7,44–46). The thresholds were additionally informed by a systematic review of dropout, attrition, and adherence patterns in culturally adapted digital mental health interventions, which identified deep cultural adaptation as consistently associated with higher adherence and lower dropout rates (47). Usability thresholds were anchored to established System Usability Scale (SUS) benchmarks (48), and satisfaction thresholds were informed by published score classifications for the measure used in this study (49).

**Table 1.** Pre-determined progression criteria.

| Criterion | Green Zone | Amber Zone | Red Zone |
| --- | --- | --- | --- |
| **Recruitment** – % of eligible students who express interest (EOI) who enrol | ≥70% | 50% - 69% | <50% |
| **Adherence** – % of active participants (those who started ≥1 module) who complete ≥1 module and click "finish" | ≥70% | 40% - 69% | <40% |
| **Retention (T1)** – % of active participants who complete the post-intervention assessment | ≥70% | 40% - 69% | <40% |
| **Retention (T2)** – % of active participants who complete the follow-up assessment | ≥60% | 40% - 59% | <40% |
| **Usability** – % of participants achieving SUS $\geq 68$ | $\geq 70\%$ | 50-69% | $< 50\%$ |
| **Satisfaction** – Mean CSQ-I score | $\geq 24$ | 20 - 23 | $< 20$ |

**Note.** SUS = System Usability Scale. CSQ-8 = Client Satisfaction Questionnaire for Internet-based intervention. T1 = post-intervention assessment. T2 = follow-up assessment. Adherence is defined as completing at least one module and selecting the "finish" button; the adherence rate is calculated among active participants (those who started $\geq 1$ module)
\*) Green = proceed; Amber = Proceed with Amendments; Red = Do not proceed without substantial revision

##### Usability

Platform usability will be assessed using the Indonesian version of the System Usability Scale (50,51), which has previously been adapted and validated in the Indonesian context. The SUS consists of 10 items rated on a 5-point Likert scale, generating a total score from 0 to 100. Scores ≥68 are conventionally interpreted as indicative of acceptable usability, while scores ≥80.3 reflect excellent usability (48). The proportion of participants achieving a SUS score of ≥68 will be used as the primary benchmark for the progression criteria. Mean SUS scores and standard deviations will also be reported descriptively to facilitate comparison with previous studies.

##### Satisfaction

Participants’ satisfaction will be measured using the Client Satisfaction Questionnaire– Internet-based Intervention version (CSQ-I) (49,52). As no validated Indonesian version was available, the CSQ-I underwent a structured translation and cultural adaptation process prior to use in this study, including forward translation, backward translation, and expert review to ensure linguistic and conceptual equivalence. The internal consistency of the adapted CSQ-I will be explored using Cronbach’s alpha. The CSQ-I consists of eight items rated on a 4-point scale, with total scores ranging from 8 to 32. Higher scores indicate greater satisfaction. A mean total score ≥24 will be considered indicative of satisfactory acceptability.

*Adherence* will be operationally defined as completing at least one intervention module, clicking the “finish” button to indicate that the participant perceived their intervention needs to have been met and considered their use of the intervention complete. The overall adherence rate will be calculated as the proportion of active participants who complete at least one module and click the “Finish” button. The overall adherence rate will be calculated as the proportion of active participants (i.e., those who started at least one module) meeting this definition. Additional engagement metrics will be collected from the platform, including the total number of modules completed, time spent on each module, number of coaching exchanges initiated, and number of logins during the six-week intervention period. To provide a more detailed descriptive profile of intervention engagement, participants will be categorised as: (1) complete users—participants who completed all 12 modules and clicked the “Finish” button; (2) partial users—participants who completed between 2 and 11 modules and clicked the “Finish” button; (3) minimum adherers—participants who completed exactly one module and clicked the “Finish” button; and (4) non-adherers—participants who did not meet the adherence criterion (i.e., completion of at least one module and clicking the “Finish” button).

*Retention* will be evaluated separately for post-intervention (T1) and follow-up (T2). Post-intervention retention will be defined as the proportion of active participants who complete the T1 assessment. Follow-up retention will be defined as the proportion who complete T2. Reasons for dropout or discontinuation will be recorded where provided and will be reported descriptively.

### Secondary Feasibility Outcomes

Secondary outcomes will examine preliminary changes across T0, T1, and T2. Consistent with the original LifeHack intervention framework and its promotive orientation, the primary targets of change are resilience and positive mental health rather than symptom reduction. Psychological distress and coping are assessed as downstream outcomes. The response and completion rate of each measure will be reported to inform the feasibility of administering these instruments in a future trial.

*Resilience* will be assessed using the Indonesian version of the Resilience Evaluation Scale (RES) (53), which measures appraisal-based resilience components including self-confidence and self-efficacy. The RES consists of 9 items rated on a 5-point Likert scale (0 = completely disagree to 4 = completely agree), with total scores ranging from 0-36. Higher scores indicate greater resilience. The Indonesian version has demonstrated acceptable internal consistency (α = 0.74–0.82).

*Positive mental health* will be measured using the Indonesian version of the Mental Health Continuum– Short Form(MHC-SF) (54,55). The MHC-SF consists of 14 items assessing emotional well-being (3 items), psychological well-being (6 items), and social well-being (5 items), rated on a 6-point Likert scale ranging from 0 (never) to 5 (every day). Total scores range from 0 to 70; higher scores indicate greater positive mental health. The Indonesian version has demonstrated acceptable reliability and validity (α = 0.75– 0.85).

#### Depression

Depressive symptoms will be assessed using the Indonesian version of the Patient Health Questionnaire–9 (PHQ-9) (56), for which acceptable reliability and predictive validity have been demonstrated in an Indonesian sample (57). The PHQ-9 comprises 9 items rated over the past two weeks on a 4-point scale (0 = not at all to 3 = nearly every day), yielding a total score from 0 to 27, with higher scores indicating greater symptom severity (minimal 0–4; mild 5–9; moderate 10–14; moderately severe 15–19; severe 20–27). In this study the PHQ-9 serves both as a screening instrument (see Recruitment) and as a secondary outcome administered at T0, T1, and T2.

*Anxiety* symptoms will be assessed using the Indonesian version of the Generalized Anxiety Disorder–7 (GAD-7) (58), for which acceptable reliability and predictive validity have been demonstrated in an Indonesian sample (57). The GAD-7 comprises 7 items rated over the past two weeks on a 4-point scale (0 = *not at all* to 3 = *nearly every day*), yielding a total score from 0 to 21, with higher scores indicating greater symptom severity (minimal 0–4; mild 5–9; moderate 10–14; severe 15–21).

*Coping behavior* will be assessed using an Indonesian-adapted version of the Coping Index (59). The instrument was previously translated and culturally adapted by the research team following standard cross-cultural adaptation procedures. Pilot testing demonstrated acceptable internal consistency, with Cronbach’s alpha coefficients of 0.68 for healthy coping and 0.72 for unhealthy coping (*unpublished data*).

### Safety Outcome

Potential negative effects associated with participation in the intervention will be assessed at T1 using the Negative Effects Questionnaire (NEQ) (60,61). The present study will use the 20-item version of NEQ. It employs a three-step conditional response format: for each of the 20 items, participants first indicate whether they have experienced the described effect (Yes/No). If they respond Yes, they then rate the severity of that effect on a 5-point Likert scale (0 = not at all distressing to 4 = extremely distressing), and subsequently indicate whether they attribute the effect to the intervention itself or to other causes. As no validated Indonesian version was available, the NEQ underwent a structured translation and cultural adaptation process prior to use in this study, including forward translation, backward translation, and expert review to ensure linguistic and conceptual equivalence. The internal consistency of the adapted version will be evaluated (Cronbach’s alpha) within the present feasibility study. Negative effects data will be reported descriptively: the proportion of participants reporting at least one negative effect, mean severity ratings among those endorsing each item, and the proportion attributing effects to the intervention will be reported across the six NEQ domains.

### Sample size

As a single-arm feasibility study, a formal statistical power calculation was not conducted, as the primary aim is to evaluate feasibility parameters — including recruitment, retention, and acceptability — rather than to test intervention efficacy. This approach is consistent with established guidance for pilot and feasibility studies, which cautions against powering such studies for efficacy outcomes (62).

Sample size was instead informed by rule-of-thumb recommendations for feasibility studies, with a minimum of 12 participants considered sufficient to estimate key feasibility parameters (63). A target of 50 enrolled participants was selected to improve the precision of preliminary estimates, provide a buffer against anticipated attrition, and remain feasible within the dual-university recruitment context. This target falls within the range supported by empirical reviews reporting a median sample size of approximately 30–60 participants in internet-based intervention feasibility studies (64). A traffic light progression criteria framework (38) was adopted to guide decision-making regarding the feasibility of proceeding to a full-scale trial, consistent with its recommended application in behavioral intervention research (65).

### Statistical analysis

All analyses will be conducted using JASP 0.95.2 (66). Descriptive statistics will be presented for all measures, including means, standard deviations, medians, and ranges for continuous variables, and frequencies and proportions for categorical variables.

Quantitative feasibility outcomes (recruitment rate, adherence rate, retention rates, and usability scores) will be calculated as proportions and compared against the predetermined traffic light progression criteria (Table 1). Usability progression criteria will be evaluated based on the proportion of participants achieving the predefined SUS benchmark (≥68). Mean SUS scores and standard deviations will also be reported descriptively. Satisfaction (CSQ-I) will additionally be summarized using means and standard deviations, with the proportion of participants meeting each benchmark reported. The internal consistency of the adapted CSQ-I and NEQ will be evaluated using Cronbach’s alpha coefficients in the present sample. Negative-effects (NEQ) data will be reported within an adverse-events framework, as described in the Safety Outcome section.

Changes in the secondary psychological outcomes (resilience, positive mental health, psychological distress, and coping) across T0, T1, and T2 will be examined using linear mixed-effects models, with time as a fixed effect and participant as a random intercept. This approach uses all available data under the missing-at-random (MAR) assumption via maximum likelihood estimation; accordingly, the amount and pattern of missing data will be described for each outcome and the baseline characteristics of participants with complete versus incomplete data compared to assess the plausibility of the MAR assumption, whereas mean substitution, pairwise deletion, and decision rules based on the proportion of missing data will not be used (67,68). Effect sizes (Cohen’s d) and preliminary estimates of outcome variability (standard deviations) will be derived to characterise the magnitude of change and to inform sample-size calculation for a future trial. As this is a feasibility study, these analyses are exploratory and will be interpreted with caution: results will be reported with 75%, 85%, and 95% confidence intervals, with a focus on estimation rather than null-hypothesis significance testing (69,70). Exploratory associations between feasibility indicators (e.g., module completion) and change in secondary outcomes will be examined using Pearson’s or Spearman’s correlations, as appropriate.

Responses to the open-ended questions will be analysed using conventional qualitative content analysis. The first author will independently code the responses and develop initial codes inductively from the data. Similar codes will be iteratively grouped into categories, with the coding framework and category development reviewed by the other authors to enhance the trustworthiness of the analysis. During interpretation, the categories will be considered in relation to relevant cultural adaptation domains (e.g., Bernal’s Ecological Validity Model) and key feasibility constructs, including acceptability, usability, and participant engagement, where appropriate. The qualitative findings will be used to complement and help interpret the quantitative feasibility outcomes and to identify areas for refinement of the intervention and study procedures prior to a definitive trial.

### Changes to the original protocol

The original protocol for this study (S4 File; English translation in S5 File) formed part of a doctoral research proposal reviewed by the Health Research Ethics Committee, Faculty of Medicine and Health Sciences, Universitas Jambi (Approval No. 4640/UN21.8/PT.01.04/2025). Before the enrolment of any participants, several changes were made, mainly to describe the methods in more detail.

The primary outcomes were specified as feasibility indicators with pre-specified progression criteria, and positive mental health and resilience became secondary outcomes; the usability benchmark was set at the conventional SUS score of 68 instead of 70 (48) The DASS-21 was replaced by the PHQ-9 and GAD-7, in line with the measures used in the original LifeHack trial (NCT06423417) (71). The Brief Resilience Scale (BRS) was replaced by the Resilience Evaluation Scale (RES), as a validated Indonesian version is available for the latter but not for the former (53,71). Screening was separated from the baseline assessment so that eligibility could be determined before full informed consent, and the exclusion criteria were described in more detail to protect participant safety. The follow-up assessment was rescheduled from 8 weeks after baseline to three months after T1, the most common follow-up period in digital mental health and resilience interventions (71, 72). Because of this longer interval between T1 and T2, focus group discussions after the intervention would further prolong data collection; they were therefore replaced by open-ended questions at the end of each module, which will be analysed using conventional qualitative content analysis rather than thematic analysis. As participation now extends over a longer period, compensation for internet data (quota) costs was increased from IDR 100,000 to IDR 150,000 and is transferred to participants’ e-wallets in two instalments, after T1 and after T2, rather than in a single payment. Finally, repeated measures ANOVA was replaced by linear mixed-effects models.

### Ethics

Ethical approval for this study has been obtained from the Health Research Ethics Committee, Faculty of Medicine and Health Sciences, Universitas Jambi (Approval No. 4640/UN21.8/PT.01.04/2025). All procedures will be conducted in accordance with the Declaration of Helsinki and applicable national and institutional ethical guidelines.

Eligible participants will provide fully informed electronic consent prior to completing the baseline assessment. Participation is voluntary, and participants may withdraw from the intervention or the study at any time without penalty or consequence to their academic standing. All participants will be informed that their decision to participate or withdraw will not affect their academic status or their relationship with the research team.

All data will be collected through a secure web-based platform. Participant confidentiality will be maintained by assigning unique identification codes and storing all data without direct personal identifiers. Access to study data will be restricted to named members of the research team.

As this study comprises a six-week intervention period followed by a three-month follow-up and is categorised as low research risk with a low-to-moderate level of participant vulnerability, a formal data and safety monitoring committee is not incorporated. Any substantive protocol modifications that may affect participant safety or study validity will be formally reviewed and approved by the ethics committee prior to implementation and reported transparently in the final study publication.

### Safety Monitoring and Potential Adverse Events

Participant safety will be supported through both pre-enrollment screening and ongoing monitoring throughout the intervention. Prior to enrollment, all prospective participants will complete an online screening questionnaire. Individuals who meet the predefined suicide-risk exclusion criterion or report severe depressive symptoms (PHQ-9 ≥20) will not be enrolled in the study. These individuals will be contacted via email within 48 hours, informed that they do not meet the eligibility criteria, and provided with written information about accessible professional mental health resources, including university counseling services and the national crisis helpline. (e.g., www.healing119.id or by calling 119 ext. 8).

Throughout the intervention, participant safety will be monitored through routine monitoring by the e-coach via the intervention platform. At the end of each completed module, participants will be invited to respond to two brief safety check-in questions regarding (1) whether they experienced emotional discomfort or deterioration in their well-being during the module, and (2) whether they experienced any unpleasant or unexpected experiences while using the programme. Participants responding affirmatively will be invited to provide a brief description of their experience.

All participant responses will be reviewed by the assigned e-coach as part of routine monitoring. Potential adverse events will be documented using a standardised monitoring log. Where participant responses indicate clinically significant psychological deterioration, fulfilment of the predefined suicide-risk criterion, or another concern requiring additional support, the case will be discussed with the principal investigator to determine the appropriate course of action, including referral to appropriate professional mental health services where indicated. Routine safety monitoring will complement, rather than replace, the systematic assessment of participant-reported negative effects using the Negative Effects Questionnaire (NEQ), which will be administered at T1.

### Timeline and Dissemination

Recruitment and screening will take place from 1 to 24 October 2026. Eligible participants will then be given one week to provide informed consent and complete the baseline assessment (T0). Access to the intervention will be provided on 1 November 2026, after which participants will enter a six-week intervention period. The post-intervention assessment (T1) will be administered immediately after participants indicate completion of the intervention or at the end of the six-week intervention period if they have not previously indicated completion. The follow-up assessment (T2) will be administered three months after T1, with the final T2 assessments expected by mid-March 2027. Overall data collection is expected to be completed by April 2027, with study results anticipated to be available by June 2027. Trial results will be disseminated via publications in peer-reviewed journals and presentations at national and international academic conferences. Findings will be reported regardless of the direction or magnitude of the results.

## Discussion

This protocol describes a single-arm pre–post feasibility study of LifeHack: Menjadi Mahasiswa Tangguh, a culturally adapted internet-based guided self-help promotive mental health intervention for Indonesian undergraduate students. This study will be among the first to systematically evaluate the feasibility, usability, and preliminary effects of this culturally adapted digital intervention, thereby addressing a significant gap in the evidence base.

Several methodological strengths of the present study are noteworthy. First, the present adaptation draws on an established, evidence-based program framework: the Caring Universities programs have demonstrated feasibility and preliminary efficacy in European university populations and have a documented evidence base in related populations. The current version of LifeHack: Menjadi Mahasiswa Tangguh has additionally undergone a rigorous and systematic cultural adaptation process informed by Bernal’s Ecological Validity Framework, including expert review by clinical psychologists and direct input from the target population. This enhances the likelihood that the intervention will be perceived as relevant and acceptable by Indonesian students. Second, the study employs a comprehensive assessment of feasibility indicators—including usability, satisfaction, adherence, retention, and safety—thereby providing a multidimensional picture of the intervention’s feasibility. Third, the use of predetermined traffic light progression criteria (38) ensures that decision-making about proceeding to a future RCT will be transparent, systematic, and evidence-based. Fourth, the inclusion of a broad set of secondary psychological outcomes (resilience, positive mental health, psychological distress, and coping) will generate preliminary effect estimates to inform sample size calculations for a future RCT.

A potential limitation is the single-arm design, which precludes causal attribution of any observed changes. Without a control group, it is not possible to distinguish intervention effects from the passage of time, regression to the mean, or other contemporaneous influences. This limitation is inherent to the phase of intervention development represented by the present study and will be addressed in a future RCT. A second anticipated challenge is the commonly high dropout rates observed in internet-based interventions among university students (7). To mitigate this, the study incorporates several evidence-based engagement strategies, including asynchronous personalized e-coaching, automated email reminders for participants who have not engaged with the platform, and compensation for completing assessments. Additionally, the broad eligibility criteria—encompassing a wide age range, varying levels of psychological distress, and allowing concurrent use of other mental health resources—were intentionally designed to reflect real-world implementation conditions. However, this heterogeneity may limit the interpretability of preliminary effect estimates.

Within the Heuristic Framework for Cultural Adaptation (31), the present study constitutes Stages 3 (Preliminary Adaptation Test) and 4 (Adaptation Refinement). Findings will directly inform whether further modifications to the adapted intervention are warranted prior to large-scale testing. Promising feasibility results will support progression to a fully powered randomized controlled trial. Unsatisfactory results will guide targeted refinement of the intervention content, delivery procedures, or study design prior to further evaluation.

## Data Availability

No datasets were generated or analysed during the current study. All relevant data from this study will be made available upon study completion

## Acknowledgements

The authors gratefully acknowledge the research assistants for their valuable contributions to this study. We also thank the LifeHack intervention development team within the Caring Universities consortium for granting permission to adapt the intervention for use in Indonesia. This publication charge is funded by Unpad through the Indonesian Endowment Fund for Education (LPDP) on behalf of the Indonesian Ministry of Higher Education, Science, and Technology and managed under the EQUITY Program (Contract No. 4303/B3/DT.03.08/2025 and 3927/UN6.RKT/HK.07.00/2025).

---

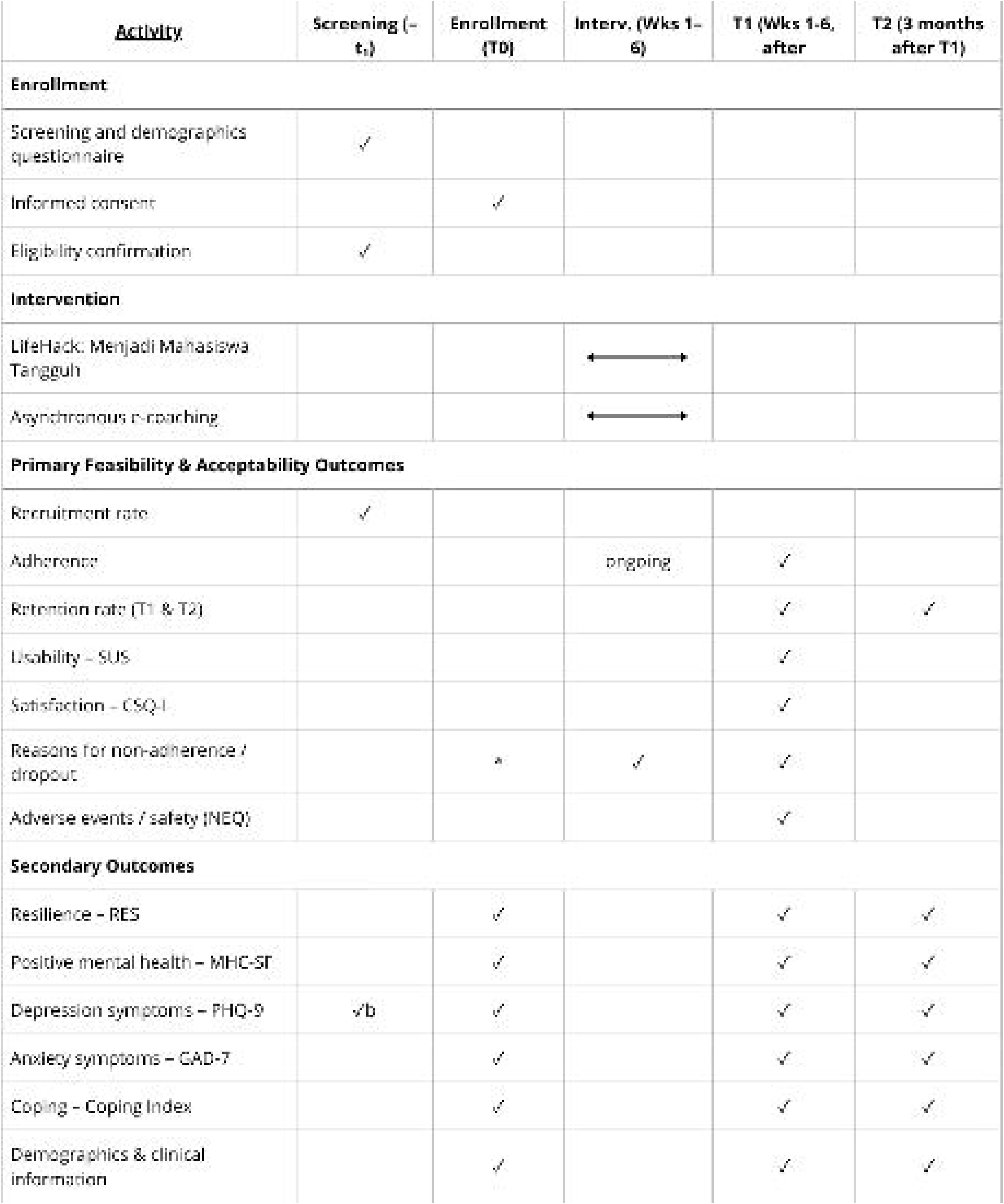

